# Association between intrahospital transfers and hospital-acquired infection among elderly patients: A retrospective case-control study in one urban UK hospital trust

**DOI:** 10.1101/2020.10.05.20191809

**Authors:** Estera Boncea, Paul Expert, Kate Honeyford, Anne Kinderlerer, Colin Mitchell, Graham S Cooke, Luca Mercuri, Céire Costelloe

## Abstract

**Background:** Intrahospital transfers have become more common as hospital staff balance patient needs with bed availability. However, this may leave patients more vulnerable to potential pathogen transmission routes via increased exposure to contaminated surfaces and contacts with individuals.

**Objective:** This study aimed to quantify the association between the number of intrahospital transfers undergone during a hospital spell and the development of a hospital-acquired infection (HAI).

**Methods:** A retrospective case-control study was conducted using data extracted from electronic health records and microbiology cultures of non-elective, medical admissions to a large urban hospital trust comprising 3 hospital sites between 2016 and 2018 (*n*=24,239). As elderly patients comprise a large proportion of hospital users and are a high-risk population for HAIs, the analysis focused on those over 65-years old. Logistic regression was conducted to obtain the odds ratio (OR) for developing a HAI as a function of intrahospital transfers until onset of HAI for cases, or hospital discharge for controls, while controlling for age, gender, time-at-risk, Elixhauser comorbidities, hospital site of admission, dominant treatment function code, intensive care admission, total number of procedures, and discharge destination.

**Results:** Of the 24,239 spells, 2879 cases were included in the analysis. 72.2% of spells contained at least one intrahospital transfer. On multivariable analysis, each additional intrahospital transfer increased the odds of acquiring a HAI by 9% (OR 1.09; 95%CI 1.05 to 1.13).

**Conclusion:** Intrahospital transfers are associated with increased odds of developing a HAI. Strategies for minimising intrahospital transfers should be considered, and further research is needed to identify unnecessary transfers. Their reduction may diminish spread of contagious pathogens in the hospital environment.

## INTRODUCTION

In recent years, pressures from an aging population coupled with inpatient bed reductions,^1^ have highlighted the importance of optimising patient flow, which encompasses patient transfers between hospitals (interhospital) and within the hospital (intrahospital). Intrahospital transfers have been variously defined in the literature as any change of location a patient undergoes within the hospital, including transfers between the emergency department (ED) and an inpatient ward, a ward and a procedure room, or two beds on the same ward.^2^ Clinical factors such as the need for a procedure or isolation due to infection may require transferring the patient.^3^ However, minimal bed availability can also induce extra intrahospital transfers.^4^ Strategies such as using empty beds in short stay-units for patients who are likely to need a longer term admission,^4,5^ or temporarily admitting patients as ‘outliers’ to clinically inappropriate wards with available beds,^6^ have been used to prevent ED congestion and meet the ‘4-hour rule’, which stipulates that patients in ED should be assessed within 4 hours of admission. A direct consequence of such strategies is that patients incur extra intrahospital transfers, with elderly patient populations disproportionately affected.^7,8^

Intrahospital transfers have been linked to a number of adverse events such as increased falls, length of stay, medication errors, delirium and hospital-acquired infections (HAIs).^2^ However, despite being established as an avenue for transmission of pathogens between hospitals,^9–11^ there is still a lack of clarity around the relationship between intrahospital patient movement and the risk of HAI.^2,12^ HAIs are defined as infections which have developed in a hospital or other healthcare delivery setting 48 hours or more following admission, or are present on day 1 or day 2 of admission in a patient discharged in the preceding 48 hours.^13^ They place a significant burden on health systems worldwide and lead to increased mortality, intensive care unit (ICU) admissions and longer hospital spells.^14^ Several factors could underlie a possible association between HAIs and intrahospital transfers. Patients with frequent transfers are exposed to a greater number of environments and contacts in the hospital, increasing their risk of coming into contact with pathogens from contaminated surfaces, other patients, or healthcare workers.^15–18^ Transferred patients may experience delays in receiving care upon admission to the new unit,^3^ thus extending their hospital stay. Increased length of stay has been associated with ward-transfers even while controlling for illness severity,^19^ and is itself a risk factor for HAIs.^20^

While some cross-sectional studies have explored the association between HAI and intrahospital transfers, these are subject to reverse causality bias, as any transfers following HAI diagnosis cannot be implicated in its cause.^19,21,22^ We identified only one case-control study considering room-transfers prior to infection alone. The group reported that for each additional room-transfer, the odds of becoming infected with *Clostridium difficile* (CDI) increased by 7%,^15^ and showed that cases have a more dispersed hospital coverage than controls using network analysis.^23^ Bush *et al*. also showed that units with high incoming transfer rates were statistically associated with new cases of CDI.^12^

To the best of our knowledge analyses of intrahospital transfers have not yet been linked with a broad range of microbiology data, despite the fact that many other nosocomial pathogens in addition to CDI have been linked to hospital surface contamination.^24,25^ In addition, no such analyses has been conducted using United Kingdom (UK) healthcare system data, or using information from multiple hospital sites. The present study applies statistical modelling to explore how the number of intrahospital transfers patients undergo influences the odds of developing any HAI in an urban hospital trust. As elderly patients make up a large proportion of hospital inpatients, and HAIs are more prevalent in this population, the analysis is focused on those over the age of 65.^26^

## METHODS

A retrospective case-control study was conducted using routinely collected electronic health records (EHR) and microbiology data.

### Data Sources

Individual level data of patients admitted between 2016 and 2018 to a hospital trust comprising three large hospital sites providing acute and specialist services were extracted from the electronic system used to routinely record patient health information. The data was anonymised, then accessed and analysed by researchers in a secure environment. The data has a hierarchical structure; an anonymised unique ID is given to each patient, and a second ID to each hospital spell of a patient, which begins when the patient is admitted and ends due to death or hospital discharge.^27^ Each hospital spell contains multiple consultant ‘episodes’, which signify a change of the consultant responsible for the patients care. The original EHR dataset provided 53 variables including: timestamped day of admission, discharge, ward IDs with entry and exit times, and treatment function codes (TFC), which refer to areas of clinical work based on the main speciality or sub-speciality of the healthcare professional responsible for the patient.^28^ In addition, diagnoses (International Statistical Classification of Diseases and Related Health Problems 10th Revision (ICD-10) and procedures (Office of Population Censuses and Surveys Classification of Interventions and Procedures (OPCS-4) aggregated by episode of care were included.^29^ The dataset was linked to a diagnostic microbiology dataset by hospital spell ID, which provided the sample collection time and date, site and the causative organism if present.

### Patient Population

The patient sample was restricted to non-elective patients, aged 65 and over, who had a hospital spell duration of at least 48 hours. Non-elective spells, which refer to unscheduled hospital admissions, were selected in order to create a more uniform patient population with regards to inpatient movement. In addition, patients categorised under a surgical TFC were excluded, as surgical patients experience a higher number of intrahospital transfers, are likely to be prescribed antibiotics prophylactically, and have a heightened risk of infection (see supplementary material Table 1 for TFC codes used).^30^ Patients that developed an infection in the first 48 hours post-admission were also excluded from the analysis, regardless of whether they had had a recent admission, in order to remove all likely cases of community-acquired infections. A flow chart of full exclusion criteria of study participants is provided (see supplementary material Figure 1).

**Table 1:**
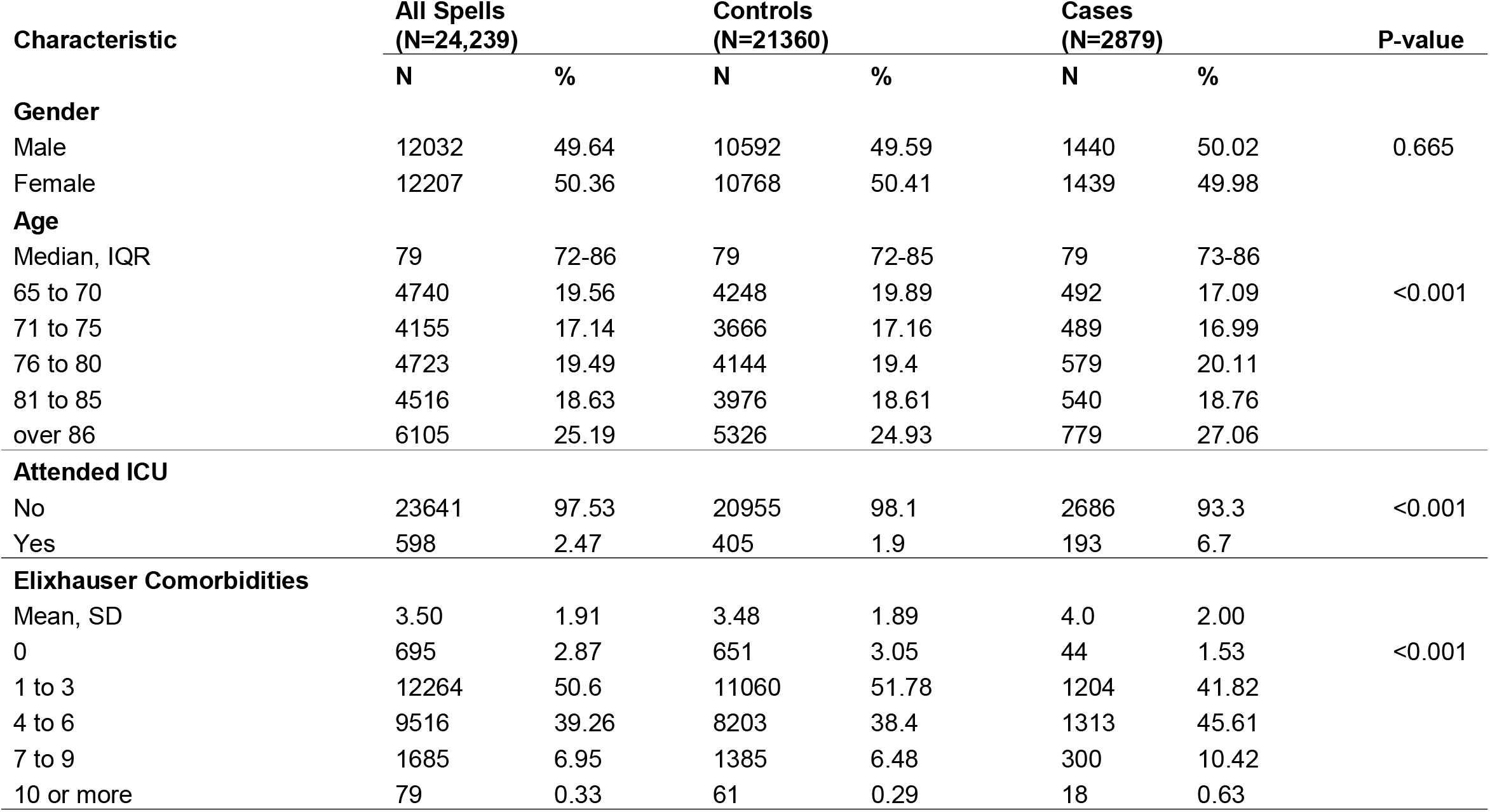
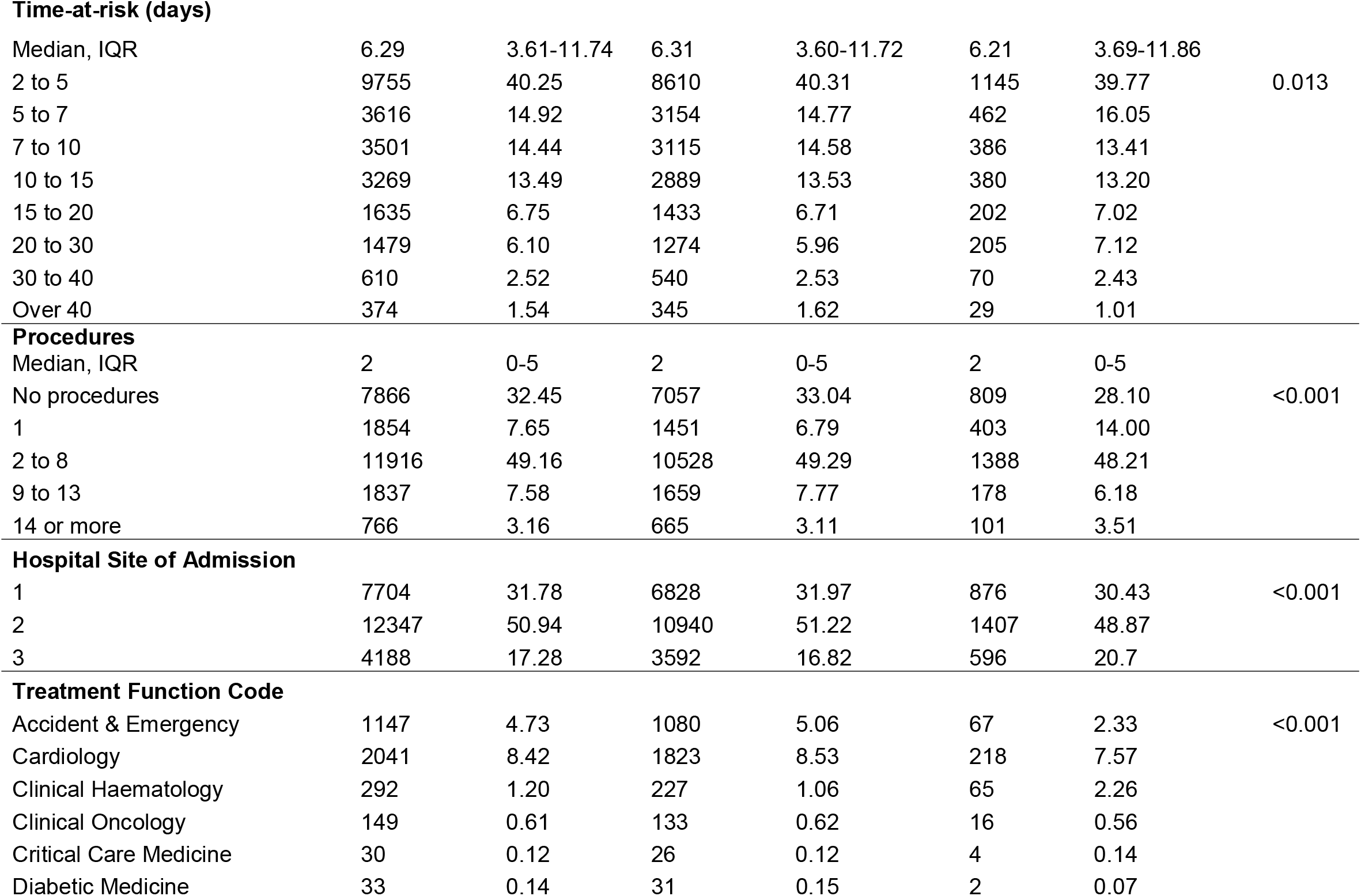
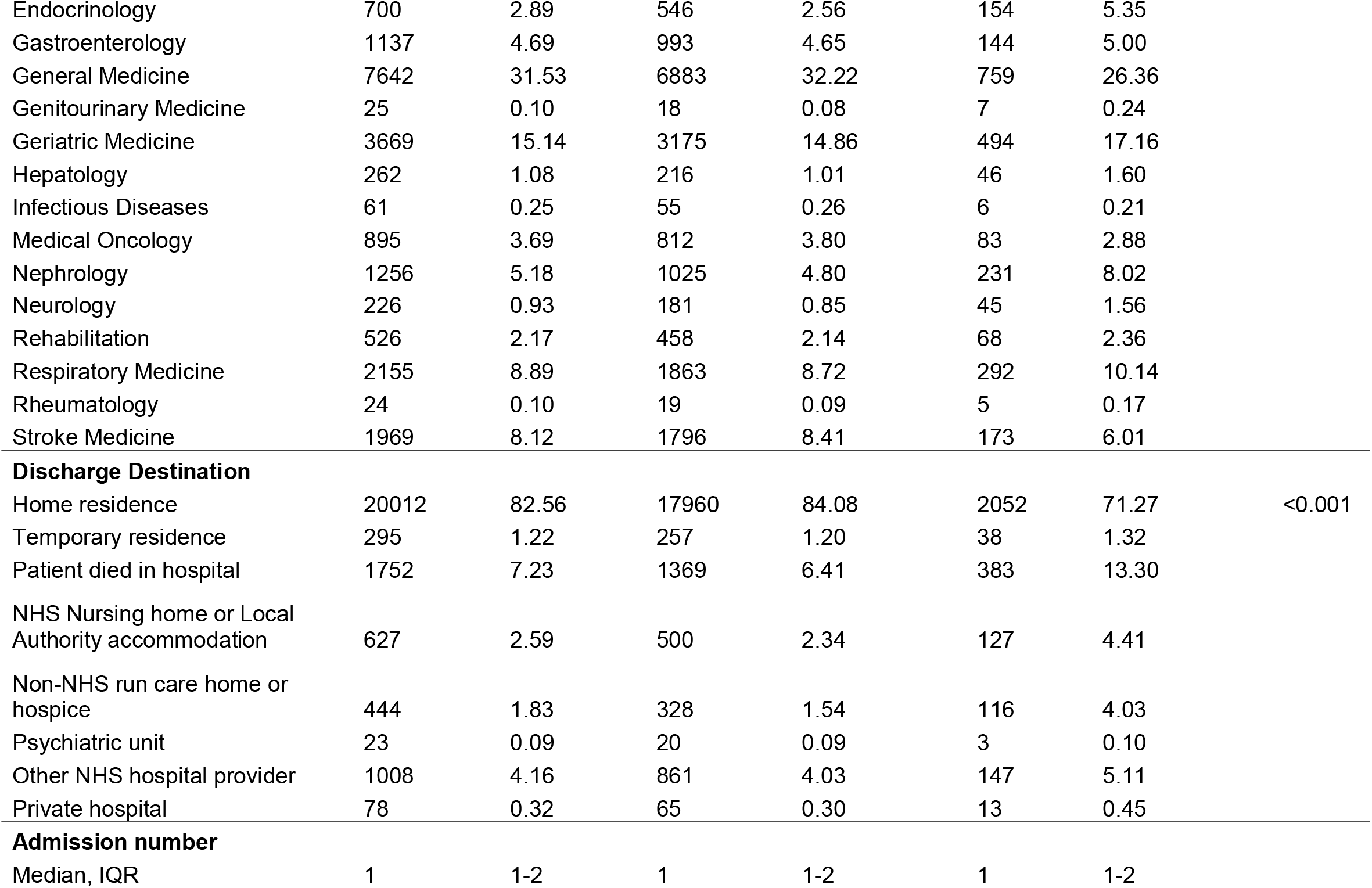
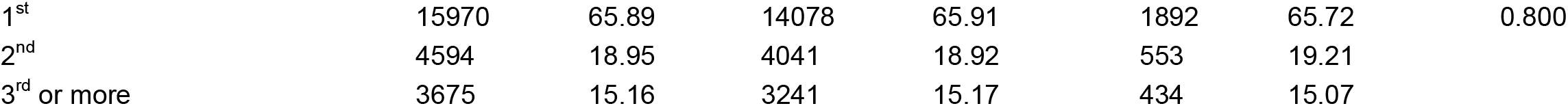
Characteristics of the 24,239 hospital spells, stratified by cases and controls. In addition, the frequency and percentage of patients across the categories of covariates used in the multivariable regression are given, with corresponding *χ*^2^ tests for significance.

**Figure 1:**
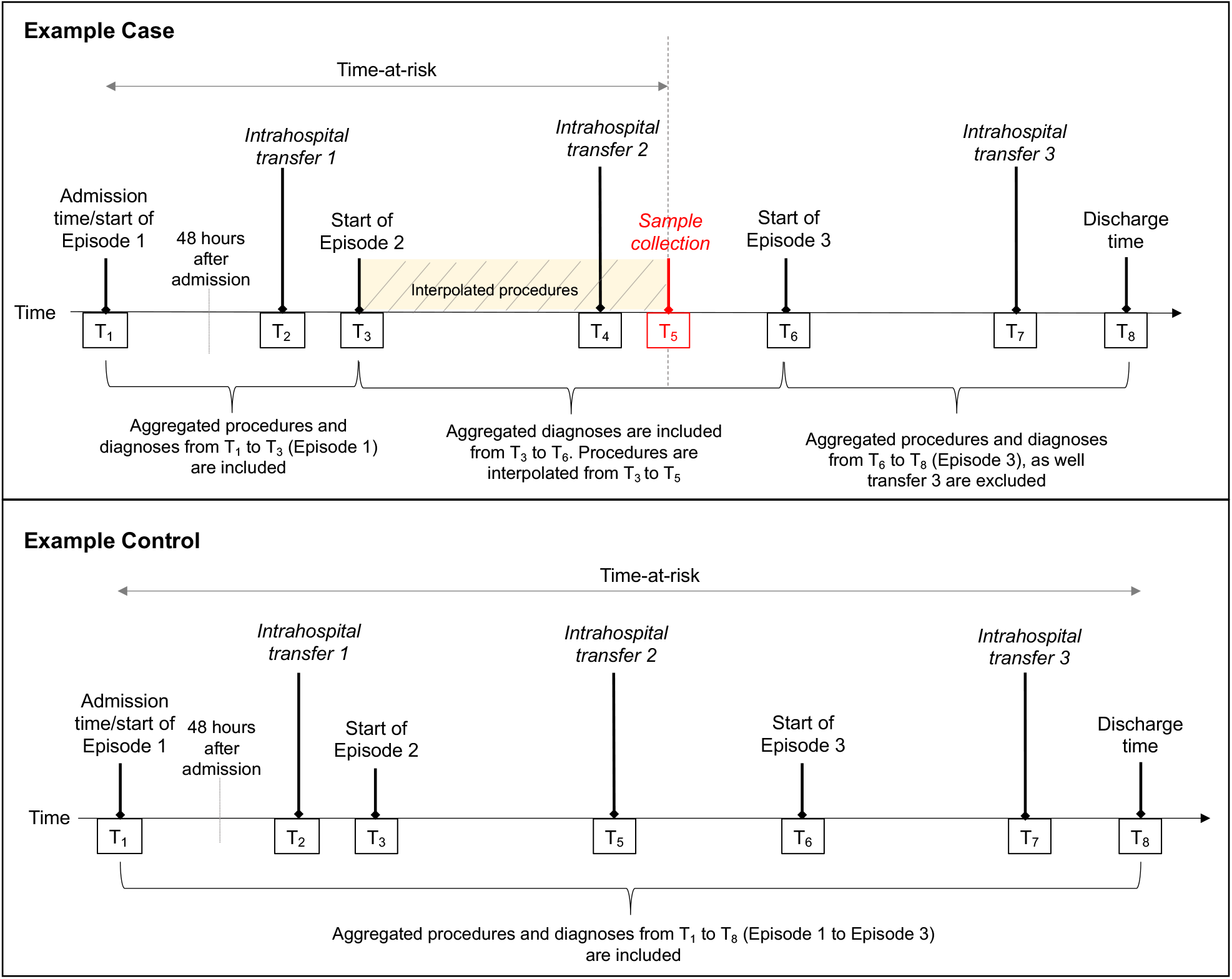
Illustration of time-at-risk definition, and time stamping in the EHR dataset using a fictitious case and control. Time-at-risk, intrahospital transfers and TFC were continuously monitored, giving a precise timestamp for their occurrence, while OPCS-4 and ICD-10 codes which are aggregated within consultant episodes. Although the optimal method of counting the OPCS-4 and ICD-10 codes for cases is to only include those which occurred from T_1_ to T_5_, due to the resolution of time stamps available in the data, only those from T_1_ to T_6_ were available. The procedure number in such episodes was interpolated between T_1_ to T_5_.

### Outcome Measure

The primary outcome was diagnosis of a HAI, defined as a positive laboratory culture (or toxin immunoassay for CDI diagnosis), collected at least 48 hours after hospitalisation. The culture sample collection date was calculated as days and fractions of days based on the time and date the sample was taken. This was used to define the time of HAI diagnosis. If a patient developed more than one infection during their hospital spell, the sample collection time and organism of their first infection was selected. Only one organism was recorded per sample, therefore co-infections were beyond the scope of this study.

### Intrahospital Transfers

The main exposure of interest was number of intrahospital transfers undergone during time-at-risk. We define time-at-risk for each spell as the length of time between a patient’s admission and the first positive culture for cases, and discharge or death for controls. All time dependent covariates were computed for the duration of time-at-risk in days and fractions of days, based on the timestamp of the variable (see Covariates). Changes in ward ID were used to derive the number of intrahospital transfers undertaken. Any change in ward ID was considered an intrahospital transfer, irrespective of the time spent in the new location, therefore capturing temporary transfers to procedure rooms or admissions and discharge lounges. Due to the granularity of the data available, bed-transfers on the same ward were not counted in our definition of intrahospital transfers. In addition, as the dataset started from inpatient admission, transfers from the ED to the first inpatient ward were not included, while all intrahospital transfers that occurred between in-hospital wards, including Ambulatory Emergency Care Centres, were used in the transfer count.

### Covariates

Elixhauser comorbidities, which comprise 31 comorbidity indicators, based on ICD-10 codes present in the episodes of care which began before the collection date were used to create a composite comorbidity measure per hospital spell, to provide comorbidity risk adjustment.^31^ Procedures were also included in risk adjustment because specific intrahospital transfers may be associated with heightened risk of infection due to the nature of the procedures that occur in the new location. For example, contamination of endoscopes and haemodialysis machines have been shown to be silent reservoirs for HAIs,^32,33^ therefore, transfers to dialysis and endoscopy procedure locations would appear to have an increased risk. OPCS-4 codes include a broad range of procedures for the diagnosis and treatment of disease, ranging from complex operations, such as transplants, to minor incisions, and non-operative procedures such as medical imaging.^29^ No classification system exists for OPCS-4 codes, therefore these were enumerated per hospital spell. All OPCS-4 codes were included from the episodes of care that began and ended *before* the sample collection date. However, for the episodes of care containing the collection date, a linear interpolation was conducted to estimate the number of procedures only up to collection date, assuming that procedures were evenly distributed over the duration of the episode (Figure 1).

Interpolated procedure number was computed by:

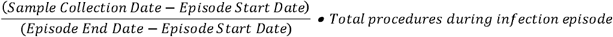

The number of procedures in this episode was interpolated independently to the episodes which ended before the collection date and the two were combined to obtain the final procedure number. While the principal diagnosis of patients was not available from the EHR data, the patient’s TFC up to collection date for cases, or hospital exit for controls, was used to categorise individuals into 20 disease related groups, and control for differences between patient groups. If multiple TFCs were assigned over a patient’s spell, the TFC under which the patient spent the longest duration was taken. Patient discharge location was also included as a categorical covariate, as discharge to a nursing home, another hospital provider or death is indicative of a frailer patient than those discharged to their own residence. Discharge destination has also been previously associated with increased ward-transfers.^19^ Lastly, as patients residing in intensive care units (ICUs) have a higher likelihood of developing a HAI,^34^ ICU admission before collection date for cases, or hospital exit for controls was included as a covariate. The final model was adjusted for age, gender, time-at-risk, Elixhauser comorbidities, hospital site of admission, dominant TFC, ICU admission, total number of procedures, and discharge destination.

### Statistical Methods

The unit of analysis in the study is the individual hospital spell. Data were initially explored descriptively to ascertain the baseline characteristics of hospital spells. Covariates which displayed a non-linear relationship with the outcome variable were grouped into categories with a similar relation to the outcome. A table of the categories chosen and numbers of observations in each category is given (Table 1). Comparisons between cases and controls were conducted using *χ*^2^ tests.

The association between intrahospital transfers and HAI was analysed using a logistic regression model. A purposeful model selection approach was taken which considered potential confounders associated with the exposure and outcome based on previous studies and clinical opinion, alongside data driven exploration. For each confounder included in the multivariable model, logistic regression model performance was assessed using goodness-of-fit tests and inspection of residuals to guide selection. No multicollinearity was found between independent variables. Patients with a time-at-risk over 57.2 days were excluded (the top 1% of time-at-risk distribution) as they are likely atypically complex cases with regards to procedures, treatment and comorbidities (see supplementary information Table 2 for results including this population). Missing data was minimal, therefore spells containing incomplete information were removed. Spells containing timing inconsistencies, such as where the final ward date did not match the discharge date were also removed.

**Table 2:** Counts and percentages of the most commonly most frequently isolated pathogens, comprising 80.9% of the 2879 cases.

| **Organism Name** | **N** | **%** |
| --- | --- | --- |
| *Clostridium difficile toxin* | 868 | 30.15 |
| *Escherichia coli* | 460 | 15.98 |
| *Pseudomonas aeruginosa* | 250 | 8.68 |
| *Enterococcus sp.* | 163 | 5.66 |
| *Klebsiella pneumoniae* | 154 | 5.35 |
| *Staphylococcus aureus* | 137 | 4.76 |
| *Coliform sp.* | 99 | 3.44 |
| *Methicillin Resistant Staph aureus* | 74 | 2.57 |
| *Coagulase negative staphylococcus* | 66 | 2.29 |
| *Enterobacter cloacae* | 57 | 1.98 |

Data exploration showed that patient-level clustering did not impact results (see supplementary material Table 3A); therefore, hospital spells from the same individual were considered independent observations, and multiple spells from one patient that met the eligibility criteria were included in the analyses. In addition, TFC-level and hospital site-level clustering was found to be minimal, and conducting a multilevel logistic regression using the command melogit in Stata V.16.0, with TFC or hospital site as a second-level cluster did not meaningfully alter the results (see supplementary material Table 3B-D).

**Table 3:** Univariable and multivariable logistic regression analysis exploring the relationship between intrahospital transfers and HAI in 24,239 hospital spells.

|  | Odds ratio for development of any HAI |  |  |  |  |  |
| --- | --- | --- | --- | --- | --- | --- |
|  | Univariable model |  |  | Multivariable model* |  |  |
|  | OR | p-value | 95% CI | OR | p-value | 95% CI |
| Intrahospital transfers | 1.08 | <0.001 | 1.05-1.11 | 1.09 | <0.001 | 1.05-1.13 |

\*Multivariable model adjusted for: age, gender, time-at-risk, Elixhauser comorbidities, hospital site of admission, dominant treatment function code (TFC), intensive care unit (ICU) admission, number of procedures and discharge destination.

### Sensitivity Analyses

It is possible that in some instances patients with a suspected infection are transferred to a side room on a different ward prior to sample collection. To assess the robustness of results against this, a sensitivity analysis was conducted with time-at-risk defined as 12 hours and 24 hours prior to the sample collection date. In this analysis the case definition was accordantly updated, meaning that cases with a time-at-risk of less than 48 hours under the new definition were removed. Finally, a second sensitivity analysis in which surgical patients were identified for exclusion by OPCS-4 codes, as opposed to TFC, was conducted to ensure that the possibility of misclassification of medical patients under a surgical TFC does not affect the results (see supplementary material Table 4A-B).

Odds ratios (OR) with 95% confidence intervals (CI) are reported for unadjusted and adjusted analyses, with a *p*-value <0.05 considered to be statistically significant. Residuals were examined to confirm the normality assumption was met. All analyses were performed using STATA version 16 software (STATA Corporation, College Station, TX) and R Studio (http://www.r-project.org).

## RESULTS

### Patient Characteristics

A total of 24,239 hospital spells were included in the analysis, pertaining to 16,018 individual patients admitted to the 3 hospital sites over the 3-year data collection period. Cases were defined as spells with a positive laboratory culture collected at least 48 hours after hospitalisation, while controls were defined as spells where the patient remained infection free for the entirety of their spell. Table 1 summarises patient characteristics by the outcome. Over the time period 11.9 % of patients developed a HAI. This is higher than the estimated HAI prevalence in acute care hospitals in England of 6.4%, based on European Point Prevalence surveys from 2016 to 2017.^35^ However, our sample included a section of the population associated with a higher risk of HAI. 49.6% of patients in the cohort were male while 50.4% were women, with a median age of 79 (IQR 72-86) and a mean of 3.5 Elixhauser comorbidities (SD 1.9).

The median time-at-risk was 6.3 days (IQR 3.6-11.7). General medicine (31.5%), Geriatric medicine (15.1%) Respiratory medicine (8.9%), Cardiology (8.4%) Stroke medicine (8.1%) and Nephrology (5.2%) comprised 77.2% of spells. Gender and admission number did not differ between cases and controls. However, patients who acquired a HAI were older than those who did not (median 79, IQR; 73-86 vs median 79, IQR; 72-85, *p*<0.001), and had a higher mean number of Elixhauser comorbidities (mean 4.0, SD 2.0 vs mean 3.5, SD 1.9; difference=0.5; *p*<0.001). Both ICU admission (6.7% vs 1.9%; difference = 4.8%; *p*<0.001), and in-hospital death (13.3% vs 6.4%, difference = 6.9%; *p*<0.001) were higher for cases. Significant differences were also found in proportions of procedures and time-at-risk intervals between cases and controls (*p*<0.001). Table 2 describes the most frequently isolated pathogens among cases.

While 27.8% of patients did not undergo any intrahospital transfers, 44.2% of patients underwent 1 intrahospital transfer during their spell, 17.1% underwent 2 transfers, and 11.0% underwent 3 or more transfers. Cases experienced more transfers than controls with 76.0% of cases undergoing at least 1 transfer, compared to 71.7% of controls. Intrahospital transfers varied marginally by TFC, with Cardiology patients moving most frequently (Median 2, IQR 1-2). Figure 2 depicts box-and-whisker plots with probability densities of intrahospital transfers by TFC.

**Figure 2:**
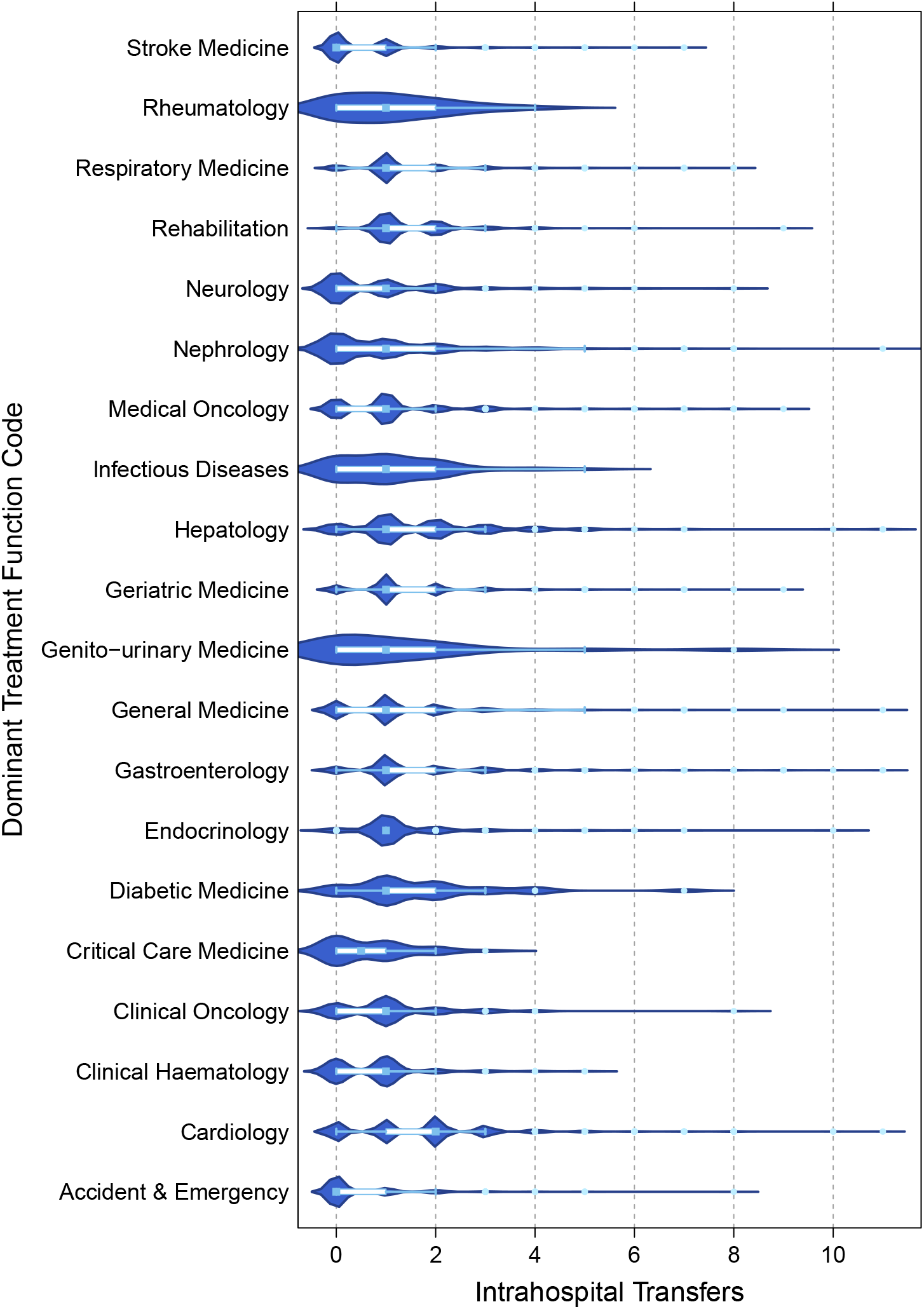
Violin and box-and-whisker plots of intrahospital transfers by the dominant TFC the patient was listed under. The length of the box represents the interquartile range (IQR), the horizontal line in the box interior represents the median, the whiskers represent the 1.5 times the IQR of the 25th quartile or 1.5 times the IQR of the 75th quartile. The violin plot depicts the probability density for each TFC group at a given ward transfer value.

Univariable logistic regressions were used to explore the effects of possible covariates on the outcome (see supplementary material Table 5) and showed that ethnicity, weekend admission and admission number were not significantly associated with the odds of developing a HAI and were excluded from the final multivariable model. Weekend admission was also modelled as an interaction in the multivariable regression model, but was not statistically significant. All other covariates included showed significant relationships, with the exception of gender, which was defined *a priori* as a covariate.

In the multivariable logistic regression results, it was found that each additional intrahospital transfer was associated with an 9% increase in the odds of developing a HAI (OR=1.09; 95%CI 1.05 to 1.13). Table 3 shows results of the univariable and multivariable logistic regression analysis (the full model is shown in supplementary material Table 6).

### Sensitivity Analysis

The effect estimate remained closely aligned to those of the main analysis when an earlier definition of time-at-risk was used. Defining time-at-risk as 12 hours prior to the collection date resulted in an OR of 1.07 (95%CI 1.03 to 1.11), which remained consistent while defining time-at-risk as 24 hours prior to the collection (OR 1.07 95% CI 1.02 to 1.11).

Exclusion of surgical patients by OPCS-4 code as opposed to TFC yielded an OR of 1.10 (95%CI 1.06 to1.13) (see supplementary material Table 4A-B).

## DISCUSSION

The present study demonstrates a robust association between the number of intrahospital transfers and the development of a healthcare associated infection (HAI), with each additional transfer increasing the odds of developing a HAI by 9% in elderly patients. We believe this is the first study to examine this association using transfer data from multiple hospital sites, as well as microbiology data from more than one organism after accounting for age, gender, time-at-risk, Elixhauser comorbidities, hospital site of admission, dominant TFC, ICU admission, procedures and discharge destination. The study contributes to a small number of studies exploring the association between intrahospital transfers and infection in an analysis which considers the chronology of events, and is concordant with other results. Our findings suggest that inpatient movement is an important risk factor for HAIs, and the decision to move a patient should be carefully considered with regards to infection risk. The use of routinely collected EHR data makes the analysis scalable, efficient and easily replicable in different settings.

The magnitude of the association between the odds of developing a HAI with each additional intrahospital transfer is similar to previously reported values by McHaney-Lindstrom *et al*., who used a similar time-at-risk approach.^15^ The group conducted nearest-neighbour matching on the admitting department to achieve a homogenous distribution of patient health conditions. However, number of procedures can vary widely between patients in the same department. This is crucial to control for, as intrahospital transfers to areas where invasive procedures occur may confound the risk from the procedure with the risk of the transfer to the procedure room. Our study used a conservative approach to adjust for interventions by including all OPCS-4 codes recorded up to infection diagnosis, and shows that these do not fully explain odds of acquiring an infection. Cross-sectional studies have reported larger effects, such as a 1.59 increase in odds of developing a HAI for one ward-transfer compared to no transfers,^19^ and a 1.28 increase in odds of developing a wound infection for each additional ward-transfer.^21^ This discrepancy likely results from the fact that cross-sectional studies do not demarcate between transfers that occurred before infection and those that occurred after infection. In a univariable analysis using intrahospital transfers for the entire hospital spell in both cases and controls, our data also showed a much larger effect, with the odds of developing a HAI increasing by 47% with each additional intrahospital transfer (OR=1.48; 95%CI 1.44 to 1.51).

While a small number of studies have previously explored the association between intrahospital transfers and HAI directly, the underlying hypothesis which implicates transfers in the horizontal transmission patterns of HAIs is in-line with results from several other study designs, providing insight into possible mechanisms. Although findings have been mixed, single-patient rooms are thought to reduce pathogen transmission opportunities and the incidence of HAIs through the hypothesised mechanism of reducing person-to-person contact and person-surface-person contacts.^16,36–38^ As intrahospital transfers increase both mixing of patients and exposure to hospital surfaces, reducing the number of non-essential transfers may be a comparable intervention, but require less resources. Intrahospital transfers also increase the individual’s exposure to other patients in the hospital. Interestingly, similar results have been reported with regard to the number of total roommate exposures per day and associated risk of CDI (Hazard Ratio (HR) =1.11), MRSA (HR=1.10) and vancomycin-resistant *Enterococcus* (HR=1.11).^39^ In addition, a requirement of 1.7 nurses has been reported to conduct a transfer, and 1.9 nurses to receive one.^40^ Transferred patients therefore necessarily experience extended interactions with hospital staff. As contacts between hospital staff and patients have been shown to promote infection spread,^18,41^ reducing intrahospital transfers would also lower opportunities for cross-infection through staff-to-patient contact. Lastly, conducting transfers is a laborious process, and has been shown to be a significant, and at times unaccounted, driving factor in nursing workload.^40,42^ High workload is a known barrier to infection prevention and control practice adherence and has been shown to be a risk factor for HAI spread.^43^ The contribution of intrahospital transfers to heavy workloads may therefore increase infection transmission indirectly.

We are aware of some limitations of our study. While proxy markers were used to adjust for illness severity, the data available did not include physiological data which could be used to compute disease severity markers such as Acute Physiology and Chronic Health Evaluation (APACHE) II score to control for patient’s baseline risk for infection.^44^ Some limitations around using OPCS-4 codes also exist, such as a lack of hierarchy with regards to invasiveness of procedures, and not all minor routine procedures are included, for example urethral catherisation will only be recorded in cases of urinary retention if conducted routinely. Unavailability of information on prescription of antibiotics and proton pump inhibitors, which have been linked to development of HAIs may also result in unobserved confounding.^45,46^ In particular, the separate effects of a procedure, prophylactic antibiotic use prior to the procedure, and movement to the procedure location on risk for HAI cannot be fully deciphered without this information. However, McHaney-Lindstrom *et al*. show that including antibiotic prescription information in their analysis did not attenuate the effect between room-transfers and CDI development.^15^ Finally, EHR information cannot account for staff movement, which has been implicated in infection spread,^47–49^ or capture casual patient movement, both of which may impact infection risk. The study is also limited by factors common to all routine data-based analyses. While steps were taken to remove spells containing timestamp inconsistencies, the possibility of timestamp inaccuracies or diagnostic coding errors cannot be avoided. A prospective study could ensure these are accurately recorded, allow for the collection of specific clinical data on illness severity and consider factors such as reasons for the transfer. While the findings of this study are of relevance to elderly patients attending other NHS hospitals, they may not be generalisable to a younger, healthier population.

These findings have particularly important implications for patients at higher risk of intrahospital transfers, such as outlying patients, who may temporarily reside on a clinically inappropriate ward until they can be transferred to a bed on an appropriate ward. It is estimated 7.5% of UK inpatients are outliers, many of whom will be older and frailer.^8,50^ Use of this patient flow strategy should be balanced with the risk accompanying moving a patient and avoided in individuals who are highly susceptible to infection. Increasing use of portable diagnostics, such as portable CT scanners, to prevent transfers to procedure wards may provide a timely solution to minimising patient movement.^51^ Reducing intrahospital transfers may not only lower the baseline levels of pathogen transmission within a hospital, but prove critical in the current COVID-19 outbreak, where patient movement could increase risk of hospital-acquired COVID-19.^52^

Amid widespread bed reductions, the NHS has been accommodating more patients in fewer beds. However, an unintended consequence of this may be an increase in intrahospital transfers. The present study demonstrates that, for elderly patients, each extra intrahospital transfer confers a 9% increase in the odds of developing a HAI. Further research is needed to better characterise unnecessary intrahospital transfers and consider strategies for minimising transfers. Their reduction could diminish the spread of contagious pathogens in the hospital environment and lighten workloads of staff in a stretched healthcare system.

## Data Availability

De-identified patient data cannot be made publicly available due to information governance restrictions. Access to the datasets used in this paper via a secure environment will be reviewed on request by Imperial College Healthcare NHS Trust

## ADDITIONAL INFORMATION

### Author Contributions

EB, PE and CC conceptualised the research question. EB, overseen by PE conducted the data analysis. EB wrote and revised the manuscript. CC, PE, GSC, AK and KH edited the manuscript. LM contributed to data curation and anonymisation. CC is the study guarantor.

## Acknowledgements

This research has been completed using National Institute for Health Research Health Informatics Collaborative (NIHR HIC) data resources and supported by the Imperial Clinical Analytics Research and Evaluation (iCARE) team. Data management (and analytical support) was provided by the Big Data and Analytical Unit (BDAU) at the Institute of Global Health Innovation (IGHI). The authors also thankful to Dr Charles Coughlan for his clinical advice.

