## Supplementary Material for "Association between intrahospital transfers and hospital-acquired infection among elderly patients: A retrospective case-control study in one urban UK hospital trust"

**Supplementary Tables and Figures**

Table 1: A list of the treatment function codes, from those present in the unfiltered dataset, used to select surgical patients.

| **List of Treatment Function Description used to identify surgical patients** | **Treatment Function Code** |
| --- | --- |
| Anaesthetics | 190 |
| Blood and Marrow Transplantation | 308 |
| Breast Surgery | 103 |
| Cardiac Surgery | 172 |
| Cardiothoracic Surgery | 170 |
| Colorectal Surgery | 104 |
| Ear Nose and Throat | 120 |
| General Surgery | 100 |
| Gynaecology | 502 |
| Hepatobiliary & Pancreatic Surgery | 105 |
| Neurosurgery | 400 |
| Ophthalmology | 130 |
| Oral Surgery | 140 |
| Paediatric Ophthalmology | 216 |
| Paediatric Surgery | 171 |
| Paediatric Trauma And Orthopaedics | 214 |
| Paediatric Urology | 211 |
| Pain Management | 191 |
| Plastic Surgery | 160 |
| Podiatric Surgery | 663 |
| Thoracic Surgery | 173 |
| Transplantation Surgery | 102 |
| Trauma & Orthopaedics | 110 |
| Urology | 101 |
| Vascular Surgery | 107 |
| Upper Gastrointestinal Surgery | 106 |

Table 2: Univariable and multivariable logistic regression analyses exploring the relationship between intrahospital transfers and hospital acquired infection in all time-at-risk distribution of patients (N=24,483). Multivariable model results were adjusted for: age, gender, time-at-risk, Elixhauser comorbidities, hospital of admission, dominant treatment function code (TFC), intensive care unit (ICU) admission, number of procedures and discharge destination.

|  |  | | **Odds ratio for development of any HAI** | | | | | | |
| --- | --- | --- | --- | --- | --- | --- | --- | --- | --- |
|  |  | | **Univariable model** | | | **Multivariable model** | | | |
| **Intrahospital transfers** |  | OR  1.07 | | *p*-value  <0.001 | 95% CI  1.04-1.11 | | OR  1.08 | *p-*value  <0.001 | 95% CI  1.04 – 1.12 |

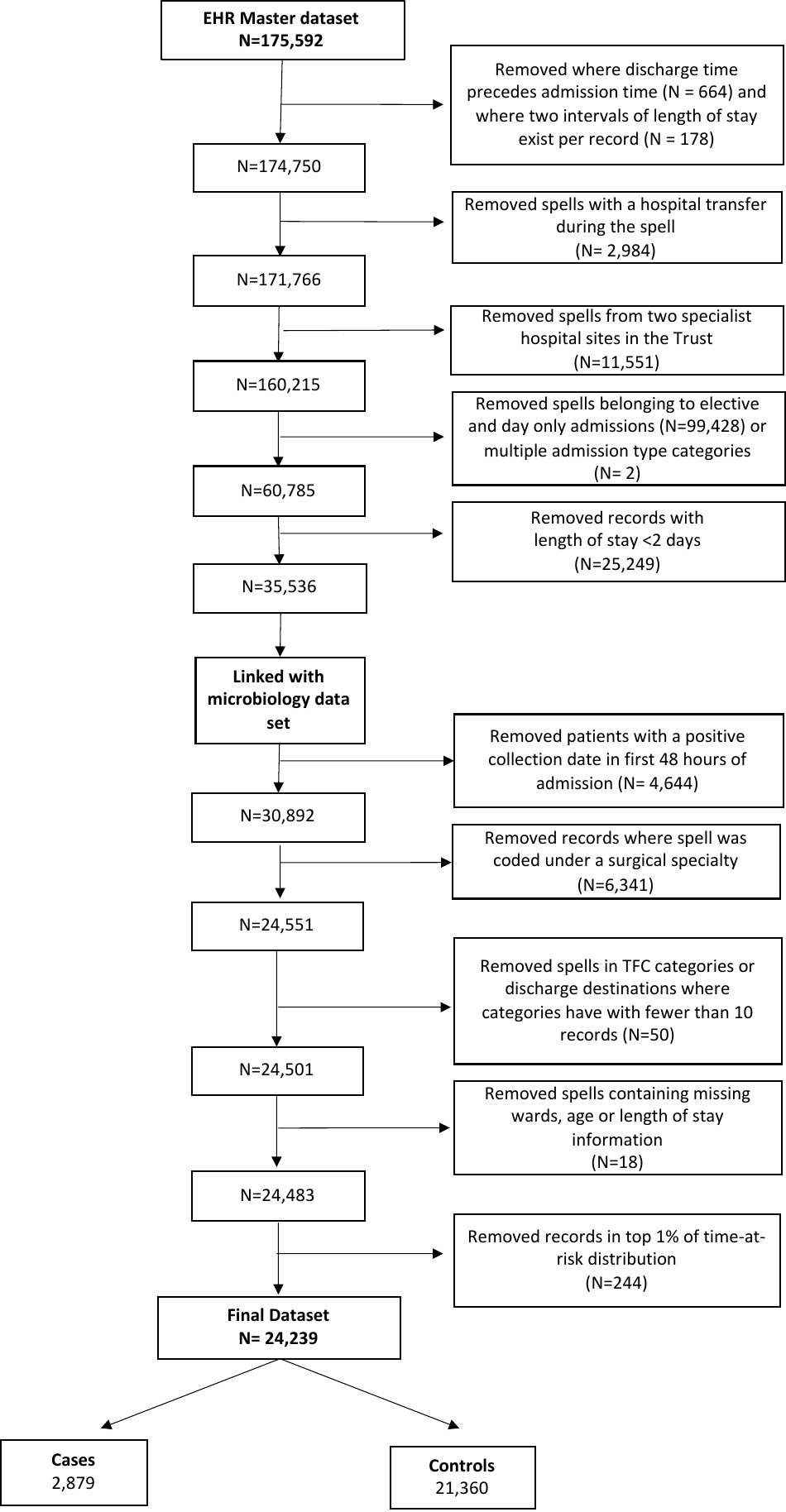

Figure 1: Flow chart depicting data linking, selection criteria and number of spells included in the analysis.

**Alternative Model Results:**

The residual intraclass correlation coefficient (ICC), which computes the proportion of variability explained by the presence of clusters was checked at the patient, treatment function code (TFC) and hospital level, and as clustering was found to be minimal a logistic regression was chosen as the final model. However, several other models were ran for data exploration. Univariable and multivariable regressions were ran using Stata’s estimation command with the vce(cluster *clustvar)* option to obtain a robust variance estimate that adjusts for within-cluster correlation at the patient level (Table 3A). In addition, univariable and multivariable logistic multilevel models (hospital spells within TFCs, and hospital spells within hospital site) were run with both random intercepts for TFCs and hospital sites, and with random slope for intrahospital transfers (Table 3B-D), using Stata’s melogit command. The model with random intercept for hospital site and random slope for intrahospital transfers did not converge. All multivariable model results were adjusted for: age, gender, time-at-risk, Elixhauser comorbidities, hospital of admission, dominant TFC, ICU admission, number of procedures and discharge destination.

Table 3A: Univariable and multivariable logistic regression results using cluster-robust standard errors for clustering at the patient-level.

|  |  | **Odds ratio for development of any HAI** | | | | | | |
| --- | --- | --- | --- | --- | --- | --- | --- | --- |
|  |  | **Univariable model** | | | **Multivariable model** | | | |
| **Intrahospital transfers** |  | OR  1.08 | *p*-value  <0.001 | 95% CI  1.05– 1.11 | | OR  1.09 | *p-*value  <0.001 | 95% CI  1.05 – 1.13 |

Table 3B: Results of multilevel univariable and multivariable model using random intercept for treatment function code (TFC).

|  |  | **Odds ratio for development of any HAI** | | | | | | |
| --- | --- | --- | --- | --- | --- | --- | --- | --- |
|  |  | **Univariable model** | | | | **Multivariable model** | | |
| **Intrahospital transfers** |  | OR  1.06 | *p*-value  <0.001 | 95% CI  1.03-1.09 | OR  1.09 | | *p-*value  <0.001 | 95% CI  1.05-1.13 |

Table 3C: Results of multilevel univariable and multivariable logistic regression a random intercept for treatment function code (TFC) and random slope for intrahospital transfers.

|  |  |  | | | **Odds ratio for development of any HAI** | | | | | | | |
| --- | --- | --- | --- | --- | --- | --- | --- | --- | --- | --- | --- | --- |
|  |  | **Univariable model** | | | | | | |  | **Multivariable model** | | |
| **Intrahospital transfers** | | |  | OR  1.06 | | *p*-value  <0.001 | 95% CI  1.00-1.11 | OR  1.09 | | | *p-*value  <0.001 | 95% CI  1.04-1.14 |

Table 3D: Results of multilevel univariable and multivariable using a random intercept for hospital site of admission

|  |  | | **Odds ratio for development of any HAI** | | | | | | |
| --- | --- | --- | --- | --- | --- | --- | --- | --- | --- |
|  |  | | **Univariable model** | | | **Multivariable model** | | | |
| **Intrahospital transfers** |  | OR  1.08 | | *p*-value  <0.001 | 95% CI  1.04-1.11 | | OR  1.08 | *p-*value  <0.001 | 95% CI  1.04 – 1.12 |

**Sensitivity analysis using time-at-risk prior to the collection date:**

Table 4A: Univariable and multivariable model results when specifying time-at-risk as 12 hours prior to sample collection date for patients (N=24,011).

|  |  | | **Odds ratio for development of any HAI** | | | | | | |
| --- | --- | --- | --- | --- | --- | --- | --- | --- | --- |
|  |  | | **Univariable model** | | | **Multivariable model** | | | |
| **Intrahospital transfers** |  | OR  1.07 | | *p*-value  <0.001 | 95% CI  1.03-1.11 | | OR  1.07 | *p-*value  <0.001 | 95% CI  1.04 – 1.11 |

Table 4B: Univariable and multivariable model results when specifying time-at-risk as 24 hours prior to sample collection date for patients (N=23,781).

|  |  | | **Odds ratio for development of any HAI** | | | | | | |
| --- | --- | --- | --- | --- | --- | --- | --- | --- | --- |
|  |  | | **Univariable model** | | | **Multivariable model** | | | |
| **Intrahospital transfers** |  | OR  1.07 | | *p*-value  <0.001 | 95% CI  1.02-1.11 | | OR  1.07 | *p-*value  <0.001 | 95% CI  1.04 – 1.11 |

**Sensitivity analysis for surgical patient’s selection strategy by OPCS-4 codes.**

While OPCS-4 intervention codes capture a wide range of procedures, they do not weigh majorly invasive procedures differently to minimally invasive procedures. They are broadly categorised into Major, Intermediate, Minor and non-operative procedures, but the individual codes themselves are not classified. The National clinical coding standard does however provide some key words associated with each of these groups. Therefore, the OPCS-4 code descriptions were searched for the terms “Total removal”, “Replacement”, “Transplant”, “Partial removal”, “Destruction” and “Reconstruction Repair” which are associated with Major and Intermediate procedures. This returned 1,250 individual codes. Spells which contained these codes at any point were removed from this analysis (N=1091) resulting in a final patient sample of 29,425.

Table 5: Univariable and multivariable logistic regression analysis exploring the relationship between intrahospital transfers and HAI in medical patients ascertained by OPCS-4 codes (N=29,425).

|  |  | **Odds ratio for development of any HAI** | | | | | | |
| --- | --- | --- | --- | --- | --- | --- | --- | --- |
|  |  | **Univariable model** | | | **Multivariable model** | | | |
| **Intrahospital transfers** |  | OR  1.09 | *p*-value  <0.001 | 95% CI  1.06-1.12 | | OR  1.10 | *p-*value  <0.001 | 95% CI  1.06-1.13 |

**Univariable Results**

Table 5: Univariable logistic regression analyses exploring the relationship between covariates and outcome of hospital acquired infection.

|  | **Odds Ratio** | ***p-*value** | **Lower 95% CI** | **Upper 95%CI** |
| --- | --- | --- | --- | --- |
| **Intrahospital Transfers** | 1.08 | <0.001 | 1.05 | 1.11 |
| **Gender** |  |  |  |  |
| Male | *Reference* |  |  |  |
| Female | 0.98 | 0.67 | 0.91 | 1.06 |
| **Age** |  |  |  |  |
| 65 to 70 | *Reference* |  |  |  |
| 71 to 75 | 1.15 | 0.04 | 1.01 | 1.32 |
| 76 to 80 | 1.21 | 0.00 | 1.06 | 1.37 |
| 81 to 85 | 1.17 | 0.02 | 1.03 | 1.33 |
| over 86 | 1.26 | 0.00 | 1.12 | 1.42 |
| **Attended ICU** |  |  |  |  |
| 0 | *Reference* |  |  |  |
| 1 | 3.72 | <0.001 | 3.12 | 4.43 |
| **Time-at-risk** |  |  |  |  |
| 2 to 5 | *Reference* |  |  |  |
| 5 to 7 | 1.10 | 0.10 | 0.98 | 1.24 |
| 7 to 10 | 0.93 | 0.26 | 0.82 | 1.05 |
| 10 to 15 | 0.99 | 0.86 | 0.87 | 1.12 |
| 15 to 20 | 1.06 | 0.48 | 0.90 | 1.24 |
| 20 to 30 | 1.21 | 0.02 | 1.03 | 1.42 |
| 30 to 40 | 0.97 | 0.85 | 0.75 | 1.26 |
| Over 40 | 0.63 | 0.02 | 0.43 | 0.93 |
| **Elixhauser Comorbidities** |  |  |  |  |
| 0 | *Reference* |  |  |  |
| 1 to 3 | 1.61 | 0.00 | 1.18 | 2.20 |
| 4 to 6 | 2.37 | <0.001 | 1.74 | 3.23 |
| 7 to 9 | 3.20 | <0.001 | 2.30 | 4.46 |
| 10 or more | 4.37 | <0.001 | 2.38 | 8.02 |
| **Procedures** |  |  |  |  |
| No procedures | *Reference* |  |  |  |
| 1 | 2.42 | <0.001 | 2.12 | 2.77 |
| 2 to 8 | 1.15 | 0.00 | 1.05 | 1.26 |
| 9 to 13 | 0.94 | 0.45 | 0.79 | 1.11 |
| 14 or more | 1.32 | 0.01 | 1.06 | 1.65 |
| **Hospital Site** |  |  |  |  |
| hospital site 1 | *Reference* |  |  |  |
| hospital site 2 | 1.00 | 0.96 | 0.92 | 1.10 |
| hospital site 3 | 1.29 | <0.001 | 1.16 | 1.45 |
| **Dominant TFC** | |  |  |  |
| Accident & Emergency | *Reference* |  |  |  |
| Cardiology | 1.93 | <0.001 | 1.45 | 2.56 |
| Clinical Haematology | 4.62 | <0.001 | 3.19 | 6.68 |
| Clinical Oncology | 1.94 | 0.02 | 1.09 | 3.44 |
| Critical Care Medicine | 2.48 | 0.10 | 0.84 | 7.31 |
| Diabetic Medicine | 1.04 | 0.96 | 0.24 | 4.44 |
| Endocrinology | 4.55 | <0.001 | 3.35 | 6.17 |
| Gastroenterology | 2.34 | <0.001 | 1.73 | 3.16 |
| General Medicine | 1.78 | <0.001 | 1.37 | 2.30 |
| Genitourinary Medicine | 6.27 | <0.001 | 2.53 | 15.53 |
| Geriatric Medicine | 2.51 | <0.001 | 1.93 | 3.27 |
| Hepatology | 3.43 | <0.001 | 2.29 | 5.14 |
| Infectious Diseases | 1.76 | 0.21 | 0.73 | 4.23 |
| Medical Oncology | 1.65 | 0.00 | 1.18 | 2.30 |
| Nephrology | 3.63 | <0.001 | 2.73 | 4.83 |
| Neurology | 4.01 | <0.001 | 2.66 | 6.03 |
| Rehabilitation | 2.39 | <0.001 | 1.68 | 3.41 |
| Respiratory Medicine | 2.53 | <0.001 | 1.92 | 3.33 |
| Rheumatology | 4.24 | 0.01 | 1.54 | 11.71 |
| Stroke Medicine | 1.55 | 0.00 | 1.16 | 2.08 |
| **Discharge Destination** |  |  |  |  |
| Home residence | *Reference* |  |  |  |
| Temporary residence | 1.29 | 0.14 | 0.92 | 1.82 |
| Patient died in hospital | 2.45 | <0.001 | 2.17 | 2.77 |
| NHS Nursing home or Local Authority accommodation | 2.22 | <0.001 | 1.82 | 2.72 |
| Non-NHS run care home or hospice | 3.10 | <0.001 | 2.49 | 3.84 |
| Psychiatric unit | 1.31 | 0.66 | 0.39 | 4.42 |
| Other NHS hospital provider | 1.49 | <0.001 | 1.25 | 1.79 |
| Private hospital | 1.75 | 0.07 | 0.96 | 3.18 |
| **Ethnic Code Description** |  |  |  |  |
| African | *Reference* |  |  |  |
| Any other Asian background | 0.94 | 0.73 | 0.68 | 1.31 |
| Any other Black background | 0.76 | 0.22 | 0.49 | 1.17 |
| Any other White background | 0.89 | 0.44 | 0.67 | 1.19 |
| Any other ethnic group | 0.92 | 0.54 | 0.69 | 1.21 |
| Any other mixed background | 0.71 | 0.35 | 0.36 | 1.44 |
| Bangladeshi | 0.91 | 0.74 | 0.51 | 1.62 |
| British | 0.99 | 0.96 | 0.77 | 1.29 |
| Caribbean | 0.99 | 0.92 | 0.73 | 1.33 |
| Chinese | 0.70 | 0.32 | 0.35 | 1.41 |
| Indian | 1.29 | 0.10 | 0.95 | 1.75 |
| Irish | 1.02 | 0.89 | 0.76 | 1.38 |
| Not known | 1.06 | 0.80 | 0.69 | 1.62 |
| Not stated | 0.83 | 0.20 | 0.62 | 1.10 |
| Pakistani | 0.97 | 0.90 | 0.61 | 1.55 |
| White and Asian | 0.36 | 0.16 | 0.08 | 1.51 |
| White and Black African | *Empty* |  |  |  |
| White and Black Caribbean | 1.21 | 0.59 | 0.61 | 2.41 |
| **Weekend Admission** |  |  |  |  |
| Weekday admission | *Reference* |  |  |  |
| Weekend admission | 0.96 | 0.33 | 0.87 | 1.05 |
| **Admission Number** |  |  |  |  |
| 1^st^ | *Reference* |  |  |  |
| 2^nd^ | 1.02 | 0.73 | 0.92 | 1.13 |
| 3^rd^ or more | 1.00 | 0.95 | 0.89 | 1.11 |

**Multivariable results:**

Table 6: Full multivariable logistic regression analysis exploring the relationship between independent variables and outcome of hospital acquired infection. Multivariable model results were adjusted for: age, gender, time-at-risk, Elixhauser comorbidities, hospital of admission, dominant treatment function code (TFC), intensive care unit (ICU) admission, number of procedures and discharge destination.

|  | Odds Ratio | *p*-value | Upper 95%CI | | Lower  95% CI | |
| --- | --- | --- | --- | --- | --- | --- |
| **Ward Transfers** | 1.09 | <0.001 | 1.05 | | | 1.13 |
| **Gender** |  |  |  | | |  |
| Male | *Reference* |  |  | | |  |
| Female | 1.02 | 0.708 | 0.94 | | | 1.10 |
| **Age** |  |  |  | | |  |
| 65 to 70 | *Reference* |  |  | | |  |
| 71 to 75 | 1.19 | 0.011 | 1.04 | | | 1.37 |
| 76 to 80 | 1.26 | 0.001 | 1.11 | | | 1.45 |
| 81 to 85 | 1.29 | <0.001 | 1.12 | | | 1.48 |
| over 86 | 1.41 | <0.001 | 1.23 | | | 1.61 |
| **Attended ICU** |  |  |  | | |  |
| No | *Reference* |  |  | | |  |
| Yes | 3.56 | <0.001 | 2.91 | | | 4.35 |
| **Time-at-risk** |  |  |  | | |  |
| 2 to 5 | *Reference* |  | |  |  |  |
| 5 to 7 | 0.87 | 0.027 | 0.77 | | | 0.98 |
| 7 to 10 | 0.65 | <0.001 | 0.57 | | | 0.74 |
| 10 to 15 | 0.61 | <0.001 | 0.53 | | | 0.70 |
| 15 to 20 | 0.56 | <0.001 | 0.47 | | | 0.67 |
| 20 to 30 | 0.56 | <0.001 | 0.47 | | | 0.68 |
| 30 to 40 | 0.41 | <0.001 | 0.31 | | | 0.55 |
| Over 40 | 0.22 | <0.001 | 0.14 | | | 0.33 |
| **Elixhauser Comorbidities** |  |  |  | | |  |
| 0 | *Reference* |  |  | | |  |
| 1 to 3 | 1.55 | 0.007 | 1.13 | | | 2.13 |
| 4 to 6 | 2.15 | <0.001 | 1.56 | | | 2.96 |
| 7 to 9 | 2.82 | <0.001 | 2.01 | | | 3.96 |
| 10 or more | 2.80 | 0.002 | 1.47 | | | 5.35 |
| **Procedures** |  |  |  | | |  |
| No procedures | *Reference* |  |  | | |  |
| 1 | 2.27 | <0.001 | 1.98 | | | 2.61 |
| 2 to 8 | 1.11 | 0.048 | 1.00 | | | 1.22 |
| 9 to 13 | 0.83 | 0.051 | 0.69 | | | 1.00 |
| 14 or more | 0.99 | 0.924 | 0.77 | | | 1.27 |
| **Hospital Site** |  |  |  | | |  |
| Hospital Site 1 | *Reference* |  |  | | |  |
| Hospital Site 2 | 1.05 | 0.354 | 0.95 | | | 1.16 |
| Hospital Site 3 | 1.36 | 0.015 | 1.06 | | | 1.74 |
| **Dominant TFC** |  |  |  | | |  |
| Accident & Emergency | *Reference* |  |  | | |  |
| Cardiology | 1.09 | 0.64 | 0.75 | | | 1.59 |
| Clinical Haematology | 4.08 | <0.001 | 2.61 | | | 6.37 |
| Clinical Oncology | 2.03 | 0.019 | 1.13 | | | 3.66 |
| Critical Care Medicine | 0.77 | 0.653 | 0.25 | | | 2.38 |
| Diabetic Medicine | 1.08 | 0.918 | 0.25 | | | 4.70 |
| Endocrinology | 4.53 | <0.001 | 3.30 | | | 6.23 |
| Gastroenterology | 2.26 | <0.001 | 1.65 | | | 3.10 |
| General Medicine | 1.46 | 0.005 | 1.12 | | | 1.90 |
| Genitourinary Medicine | 7.57 | <0.001 | 2.99 | | | 19.14 |
| Geriatric Medicine | 2.40 | <0.001 | 1.82 | | | 3.16 |
| Hepatology | 3.48 | <0.001 | 2.29 | | | 5.30 |
| Infectious Diseases | 1.33 | 0.53 | 0.54 | | | 3.29 |
| Medical Oncology | 1.56 | 0.012 | 1.10 | | | 2.21 |
| Nephrology | 2.96 | <0.001 | 2.03 | | | 4.32 |
| Neurology | 3.85 | <0.001 | 2.50 | | | 5.94 |
| Rehabilitation | 2.53 | <0.001 | 1.74 | | | 3.67 |
| Respiratory Medicine | 2.40 | <0.001 | 1.81 | | | 3.19 |
| Rheumatology | 3.55 | 0.02 | 1.23 | | | 10.31 |
| Stroke Medicine | 1.29 | 0.105 | 0.95 | | | 1.76 |
| **Discharge Location** |  |  |  | | |  |
| Home residence | *Reference* |  |  | | |  |
| Temporary residence | 1.42 | 0.054 | 0.99 | | | 2.01 |
| Patient died in hospital | 2.08 | <0.001 | 1.83 | | | 2.38 |
| NHS Nursing home or Local Authority accommodation | 2.88 | <0.001 | 2.33 | | | 3.57 |
| Non-NHS run care home or hospice | 3.73 | <0.001 | 2.96 | | | 4.69 |
| Psychiatric unit | 1.72 | 0.387 | 0.50 | | | 5.91 |
| Other NHS hospital provider | 1.88 | <0.001 | 1.53 | | | 2.30 |
| Private hospital | 2.15 | 0.016 | 1.15 | | | 4.01 |
